# Methamphetamine Use and Psychiatric Morbidity in Sri Lanka: Sociodemographic Insights from a Tertiary Care Clinic

**DOI:** 10.1101/2025.10.12.25337849

**Authors:** J Galhenage, A Isuru, M Dayabandara, R Hanwella

## Abstract

**Background and Objectives:** Methamphetamine (METH) use is rising across South and Southeast Asia, yet evidence on user characteristics and psychiatric morbidity remains limited. This study examines the sociodemographic features, psychiatric symptoms, and their associations among METH users in Sri Lanka.

**Methods:** A cross-sectional study recruited 245 consecutive patients attending a substance use disorder clinic at the National Hospital of Sri Lanka. Data were collected using a structured questionnaire, DSM-5 diagnostic criteria, and the Brief Psychiatric Rating Scale (BPRS).

**Results:** Most participants were male (97.6%), with a mean age of 31.3 years (SD = 9.7). Mean age of initiation was 29.2 years (SD = 10.0), with friends being the most common introducers (86.9%). Dependence was identified in 60.8%. Polysubstance use was frequent, with nicotine (93.1%) and heroin (73.9%) most common. Substance-induced psychosis (10.6%) and depressive disorder (4.9%) were the main psychiatric diagnoses. The mean BPRS score was 24.2 (SD = 8.0); suspiciousness (86%), hostility (82.5%), and depressive mood (61.6%) were the most prevalent symptoms. Higher BPRS scores were significantly associated with younger age, family history of mental illness, unemployment, recent METH use, and dependence. Logistic regression confirmed dependence as an independent predictor (OR = 6.8, 95% CI = 1.7–26.9) for higher BPRS values.

**Conclusion:** METH users in Sri Lanka are predominantly young males with high rates of dependence, polysubstance use, and significant psychiatric morbidity. These findings highlight the urgent need for integrated mental health and substance use interventions and provide data relevant for regional and global public health strategies.

## Introduction

Methamphetamine is the second most widely used illicit psychoactive substance globally (1). Its use is associated with a spectrum of psychotic presentations, ranging from brief intoxication-related states to persistent substance-induced psychotic disorders that may resemble schizophrenia (2). Studies conducted in the Southeast and East Asia have documented a wide array of health problem related to METH use (1). Beyond direct psychiatric effects, methamphetamine-induced psychosis contributes to social and occupational dysfunction, violence, and poor treatment outcomes(3,4). Recent evidence also suggests a bidirectional relationship between methamphetamine use and psychotic symptoms, where each can exacerbate the other.(5). In Sri Lanka, a survey of 50 methamphetamine users reported a mean age of initiation of 19 years, with common concurrent use of heroin, cannabis, alcohol, cigarettes, and other stimulants(6). The highest number of methamphetamine-related arrests (1762) was reported in the Colombo district in 2022(7). Methamphetamine use and arrests related offences have increased over the three years period since 2017 in Sri Lanka(6). Understanding the characteristics of high-risk populations is critical for prevention and intervention. Against this background, the present study aimed to describe the sociodemographic characteristics and psychiatric morbidity of methamphetamine users attending the National Hospital of Sri Lanka, and to explore the associations between these factors.

## Materials and methods

This cross-sectional descriptive study was conducted among methamphetamine users attending the Substance Use Disorder Management Clinic, outpatient psychiatry clinics, and inpatient psychiatry referrals from other wards of the National Hospital of Sri Lanka (NHSL). Patients were excluded if they had been receiving antipsychotic or antidepressant treatment for more than one month, had a pre-existing mental illness prior to methamphetamine use, or were critically unwell. The sample size, based on a prevalence of methamphetamine-induced psychosis of 15%,(8,9) was calculated to be 215, accounting for a margin of error of 5%, alpha error of 5% and a 10% of dropout rate. Data collection was carried out by the principal investigator of the study. Demographic data and substance use details were collected using a pretested interviewer-administered questionnaire. The study identified methamphetamine users in the Colombo district and conducted clinical assessments using DSM-5 diagnostic criteria for substance use disorder. Psychiatric symptoms were assessed using the Brief Psychiatric Rating Scale (BPRS), a semi-structured clinical interview tool(10). BPRS is considered a sensitive and specific measure for assessing psychiatric symptoms; one study has found a sensitivity of 71.4% and a specificity of 69.1% for conceptual disorganization(11). Its brevity and clinical practicality make it well suited for busy tertiary care settings in Sri Lanka, where comprehensive but time-efficient assessment is required(12). Major mental illnesses, including depressive disorder and substance-induced psychosis, were diagnosed through independent clinical assessment by a consultant psychiatrist, in addition to using DSM-5 diagnostic criteria. The study was conducted from September 2020 to February 2021.Socio-demographic characteristics were analysed as frequencies and percentages. Comparison of dichotomous variables was performed using Person’s Chi-squared test, and multiple logistic regression analysis was employed to identify the associated factors of methamphetamine use. A type 1 error of 0.05 was considered in all significance analyses. Statistical analysis was performed using IBM SPSS software. Ethical approval was obtained from the Institutional Ethics Review Committee of NHSL. Administrative approval was also granted by the hospital director and the Senior Medical Officer of the Outpatient Department.

### Results

Of 245 respondents, 239(97.6%) were males and 147 (60%) were single. The mean age was 31.30(SD=9.74) years, the minimum age was 15 years, and the maximum age was 63 years. The majority (173, 70.6%) were educated up to grade 11 and 108 (44.1%) had an income less than Rs. 50000 per month. Majority were from substance use disorder management clinic (196, 80%) followed by outpatient psychiatry clinic (32, 13.06%) and in patient psychiatry referrals (17,6.9%).

Among methamphetamine users, 23 (9.4%) were in job cat III (owners of businesses, sales representatives, managers, or administrators). The most common modes of referrals were by self or family members (142, 58%) followed by courts (68, 27.5%) for medical treatments. Out of 245 responders, 112 (45.7%) had a history of imprisonment. Seventy-six (31%) had a history of substance use among first degree family members and 9.8% had a family history of major mental illness. The commonest mental illness, other than methamphetamine use disorder, was newly diagnosed substance induced psychosis (26, 10.6%) followed by depressive disorder (12, 4.9%) and schizophrenia (9, 3.7%).

The majority of methamphetamine users’ resident within the Colombo district, and total of 83(33.9%) were from districts outside the Colombo. The highest number of substance users was reported from Colombo (52, 21.1%), Thimbirigasyaya (28, 11.4%) and Kolonnawa (23, 9.4%) divisional secretariat areas. Figure 1 illustrates the distribution of participated METH users within the Colombo district.

**Figure 1.**
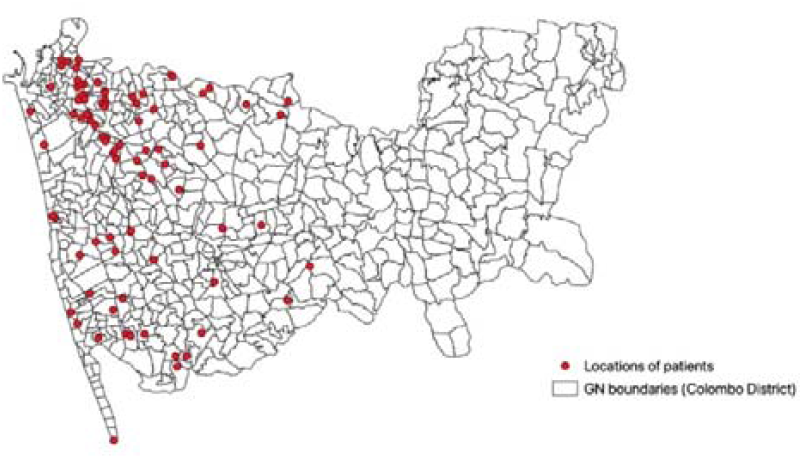
A map showing locations of participated methamphetamine users according to GN division in Colombo district (Drawn by Geographic information system)

Details of substance use characteristics are presented in Table 1. The most common concurrent drugs were nicotine, and heroin. They were used weekly or more often by 228 (93.1%) and 181 (73.9%) respectively. The next most common drugs among methamphetamine users included cannabis (116, 47.3%), pregabalin (74, 30.2%) and alcohol (26, 10.6%).

**Table 1.** Substance use characteristics among all methamphetamine(METH) users.

| **Substance use characteristic** |  | **Number (Percent)** |
| --- | --- | --- |
| Age of onset of METH use(Mean+/- SD) (Min-Max) |  | 29.16(SD=10.02)<br>(Min=14, Max=62) |
| Setting of first use | Workplace | 24(9.8%) |
|  | Friend's place | 96(39.2%) |
|  | Home or place near the home | 47(19.1%) |
|  | Party | 39(15.9%) |
|  | Prison | 6(2.4%) |
|  | Any other | 12(4.9%) |
|  | Can't remember | 8(3.3%) |
|  | School | 1(0.4%) |
|  | Rented room | 4(1.6%) |
|  | Funeral house | 8(3.3%) |
| Frequency of METH use | Daily | 39(15.9%) |
|  | Once a week or more but less than daily | 108(44.1%) |
|  | Once a month or more but less than once a week | 27(11.0%) |
|  | Less than once a month | 55(22.4%) |
| First introducer | Friend | 213(86.9%) |
|  | Workmate | 5(2.0%) |
|  | Relation / spouse/partner | 5(2.4%) |
|  | Prison inmate | 5(2.0%) |
|  | Any other | 10(4.1%) |
|  | Can't remember | 5(2.0%) |
| Last use | Within last 24 hours | 9(3.7%) |
|  | 1 day to 7 days duration | 55(22.4%) |
|  | 8 days to 30 days duration | 37(15.1%) |
|  | 1 month to 1 year duration | 141(57.6%) |
| Duration of use | Less than 30 days | 47(19.2%) |
|  | 31 days to 90 days | 13(5.3%) |
|  | 91 days to 1 year | 74(30.2%) |
|  | More than 1 year | 111(45.3%) |
| Mode of administration | Snorting by heating on a glass plate | 241(98.4%) |
|  | Smoking through a bong | 3(1.2%) |
|  | Aluminium coil and cylinder | 1(0.4%) |
|  | Intravenous | 0(0.00%) |
|  | Oral | 0 (0.00%) |
| Source of drug | Illegal drug market | 138(56.3%) |
|  | friends or relatives | 100(40.8%) |
|  | Any other | 4(1.6%) |
|  | Not willing to reveal | 1(0.4%) |
| Most commonly METH use place | Home or nearby place | 71(29%) |
|  | Friends place | 124(50.6%) |
|  | Hotel/Restaurant/Dance hall | 10(4.1%) |
|  | Work place | 10(4.1%) |
|  | Funeral houses | 1(0.4%) |
|  | Three wheel/car | 2(0.8%) |
|  | Prison | 1(0.4%) |
|  | Any other place | 14(5.7%) |
|  | Not willing to reveal | 3(1.2%) |
| DSM 5 criteria for METH dependence(At least 2 over last 12 months) | Yes | 149(60.8%) |
|  | No | 96(39.2%) |
| Concurrent use of other drugs | Yes | 234(95.5%) |
|  | No | 10(4.1%) |

The mean BPRS score was 24.25 (SD=8.00) among 245 methamphetamine users. The minimum BPRS score was 1.00 & the maximum was 49.00. The majority had scored a BPRS score of 18.00. Methamphetamine users were divided into two groups: those above and those below the mean of BPRS score. Methamphetamine users with above-mean BPRS scores were analysed further, as they exhibited more psychiatric symptoms than the other group. The line chart shows that distribution of psychotic symptoms among Methamphetamine users with above-mean BPRS scores differs from that of below-mean BPRS scorers. The most common psychotic symptoms among individuals with above-mean scored individuals were somatic concerns (2, 2.093 (SD=1.6457)), anxiety symptoms (1.96, SD=1.14), grandiosity (1.75, (SD=1.02)), depressed mood (2.7, SD=1.67), hostility (3.06, SD=1.31), suspiciousness (3.87, SD=1.75) and hallucinating behaviour (2.84, SD=1.88) (BPRS items 1,2,8,9,10,11 and 12) (Figure 2).

**Figure 2.**
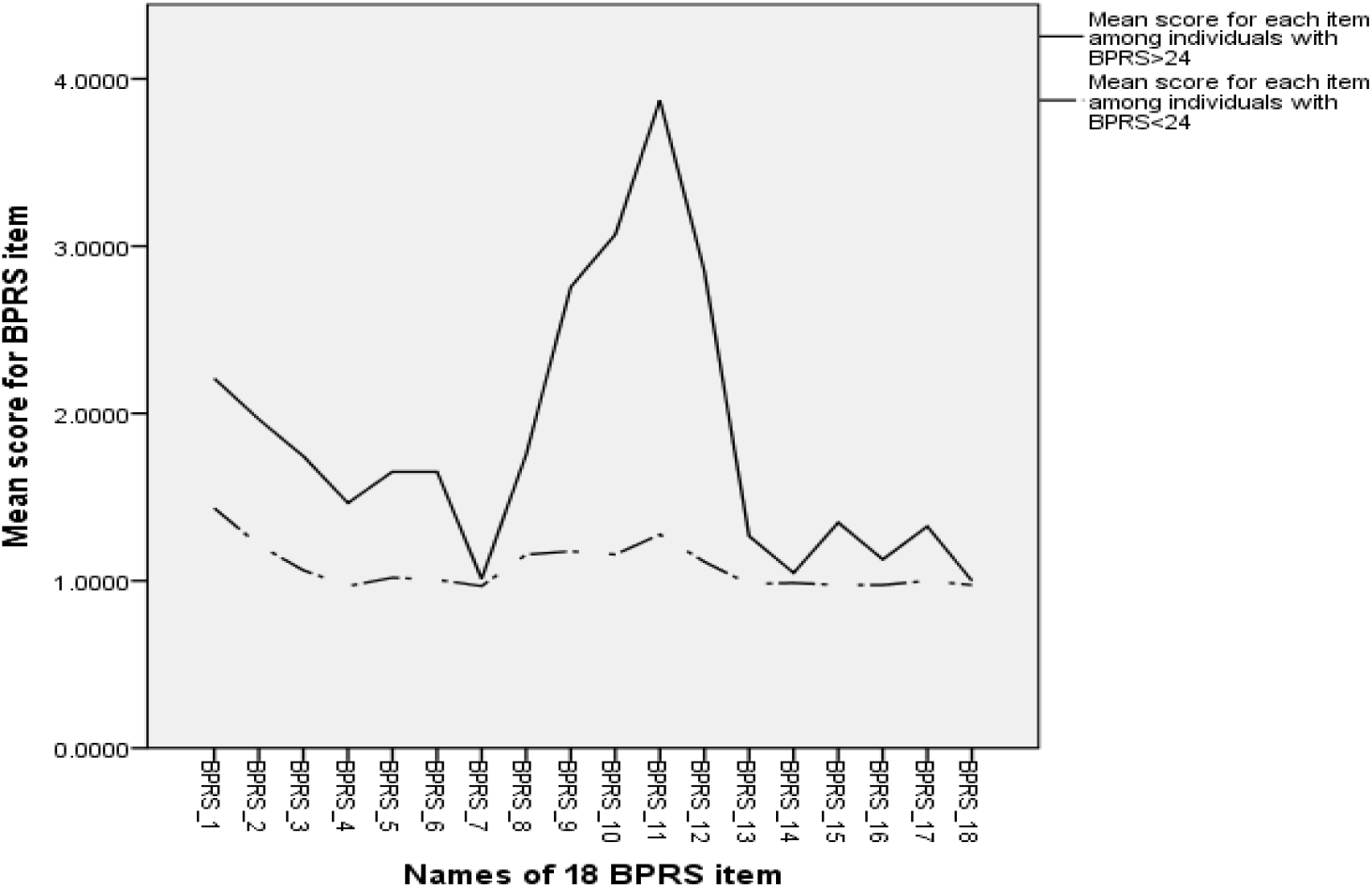
Line charts of mean scores for BPRS items of Methamphetamine users with above (continuous line) and below mean (<24) (long dashed line) total BPRS score.

Analysis of socio-demographic variables with BPRS revealed several significant associations and all associations were statistically significant (Table 2). Logistic regression revealed that methamphetamine dependence was significantly associated with psychosis (OR=6.797, 95% CI=1.71-26.87) (Table 3).

**Table 2.**
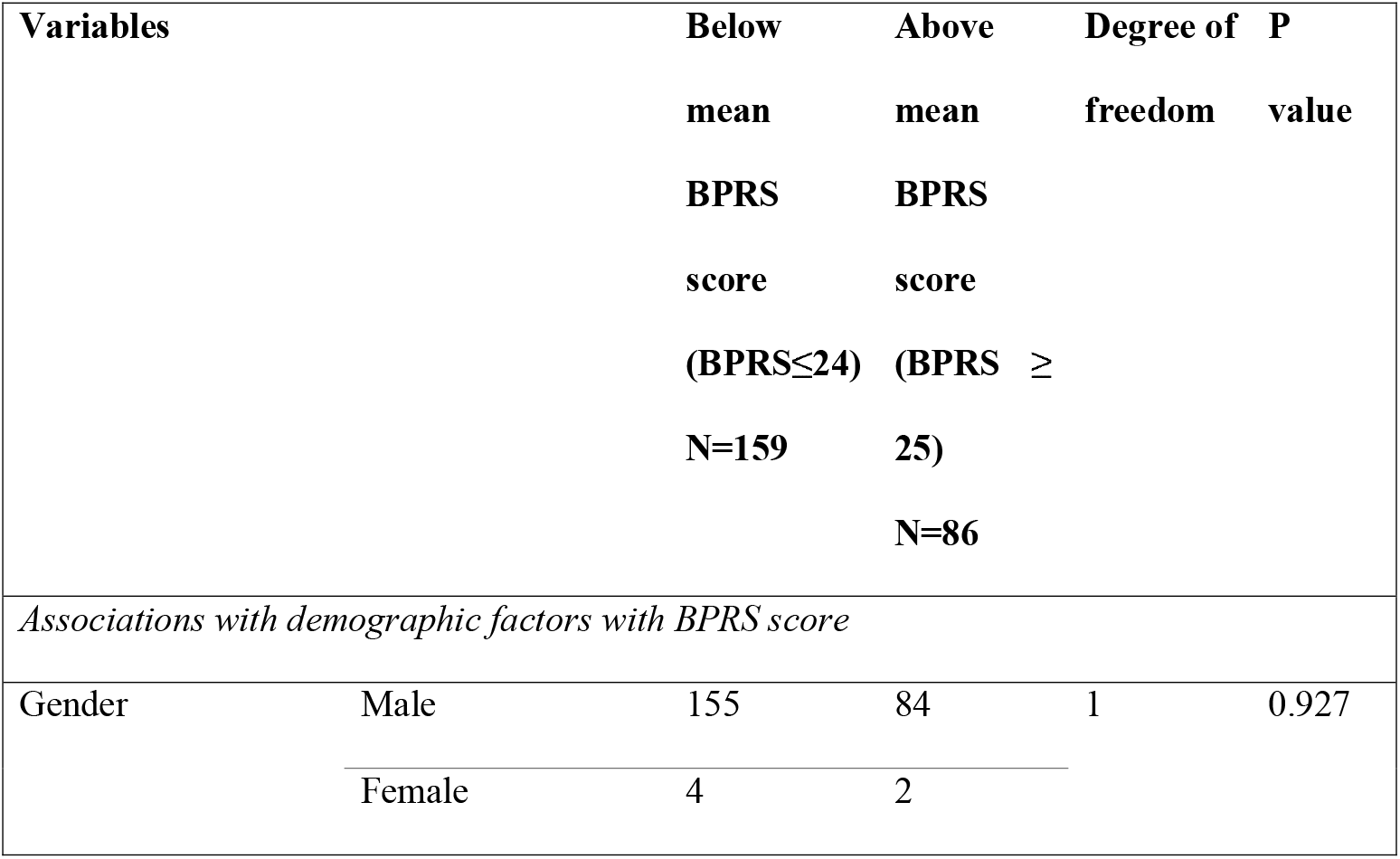
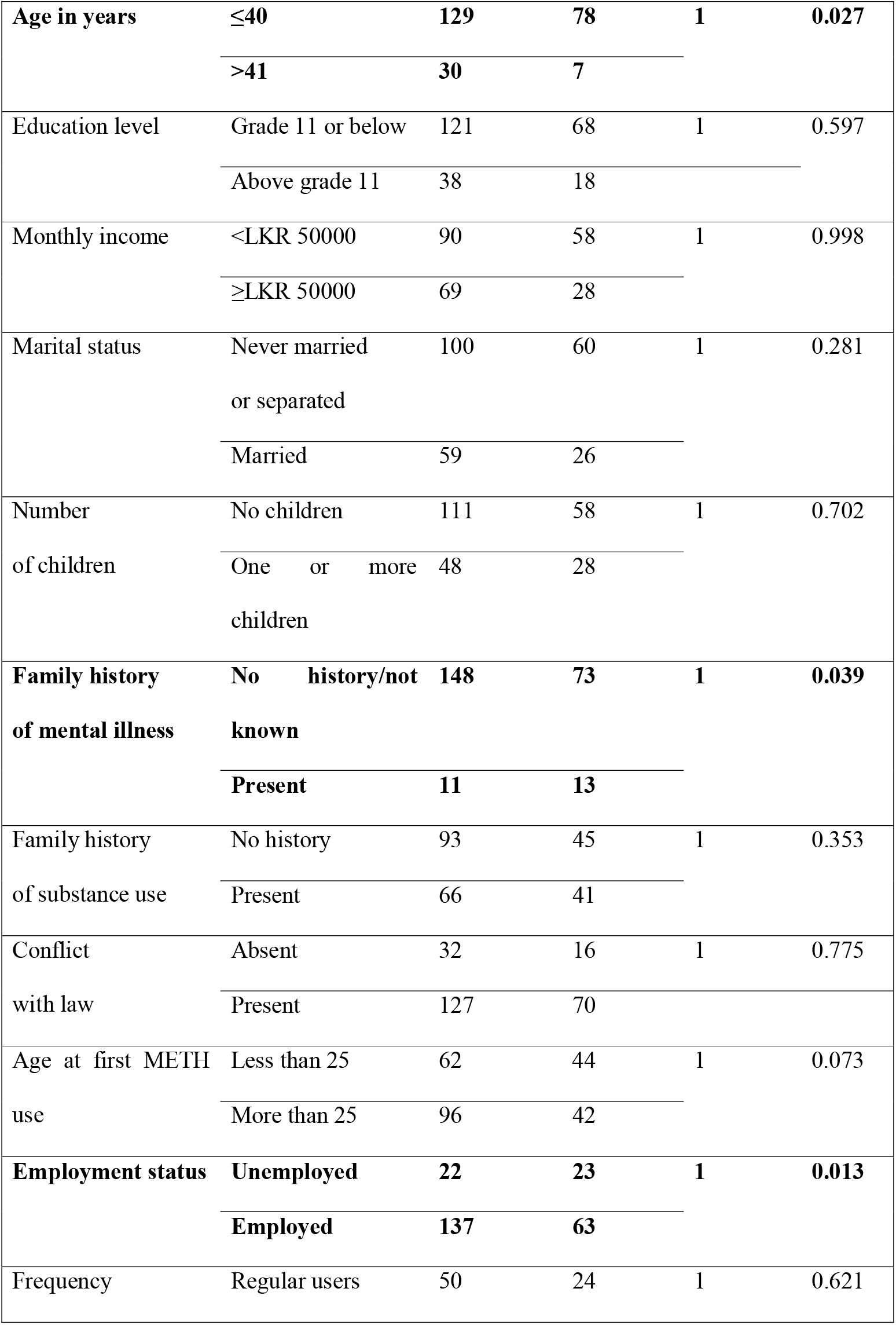
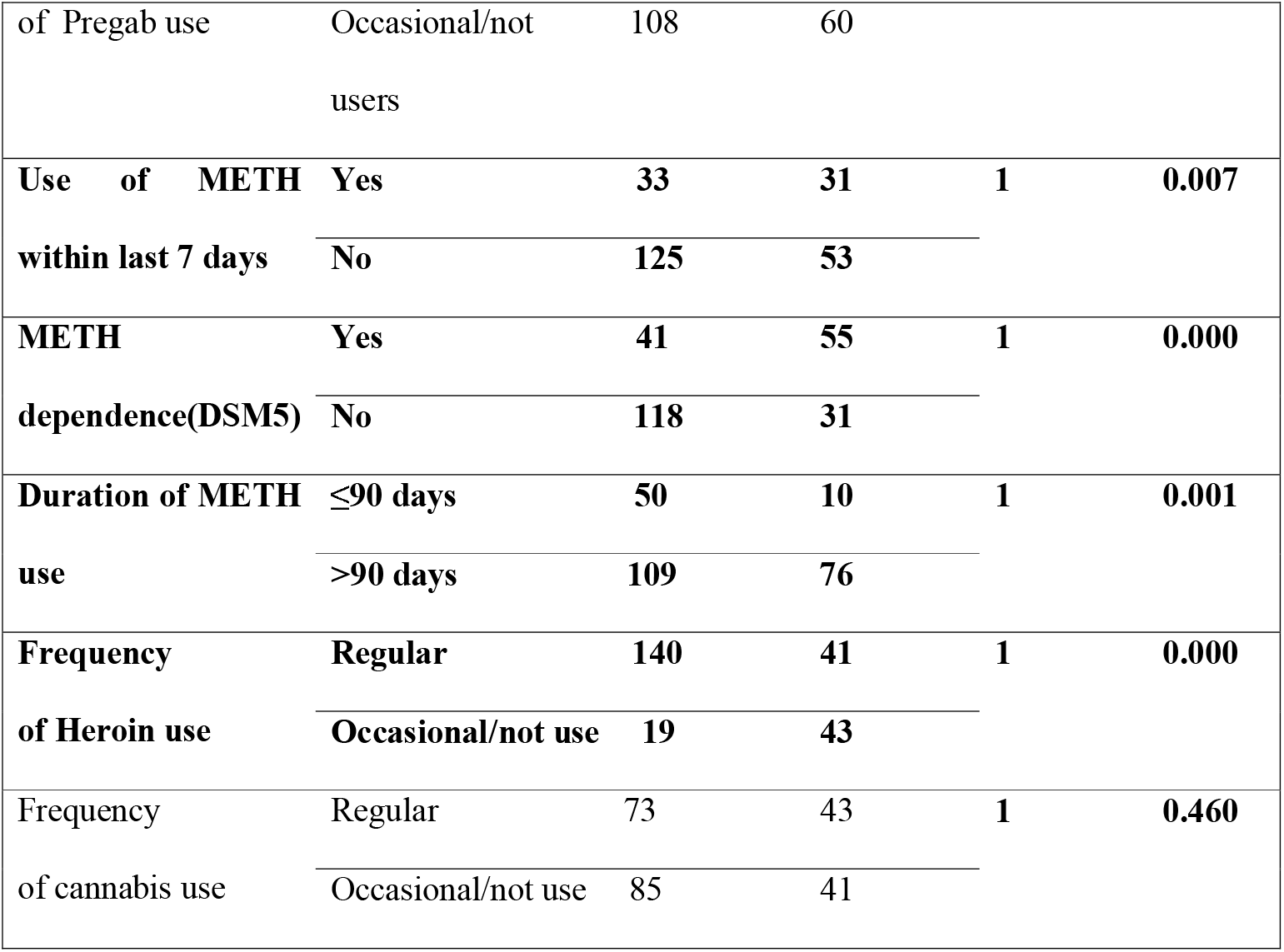
Distribution of demographic and substance use characteristics with BPRS scores among methamphetamine (METH) users.

**Table 3.** Factors associated with psychosis among methamphetamine (METH) users-logistic regression examining different character groups.

| **Character groups (variables)** | **Co efficient** | **Level of significance** | **Odds ratio** | **95% Confidence interval** |
| --- | --- | --- | --- | --- |
| Gender | 0.529 | 0.729 | 1.697 | 0.085-33.761 |
| Age | -0.312 | 0.062 | 0.732 | 0.528-1.016 |
| **Marital status** | **-4.220** | **0.012** | **0.015** | **0.001-0.399** |
| Number of children | 0.717 | 0.219 | 2.049 | 0.652-6.437 |
| Educational status | 0.281 | 0.689 | 1.324 | 0.336-5.224 |
| Monthly income | -0.224 | 0.803 | 0.799 | 0.138-4.629 |
| Employment category | 0.313 | 0.588 | 1.367 | 0.442-4.231 |
| Past conflict with law | -1.259 | 0.059 | 0.284 | 0.077-1.047 |
| Family history of substance use | -0.512 | 0.651 | 0.599 | 0.065-5.523 |
| Frequency of METH use | -0.436 | 0.263 | 0.647 | 0.301-1.388 |
| Last METH use | -0.090 | 0.906 | 0.914 | 0.204-4.101 |
| Mode of administration | -0.174 | 0.801 | 0.840 | 0.216-3.267 |
| Duration of METH use | 0.936 | 0.136 | 2.550 | 0.745-8.734 |
| **METH dependence** | **1.917** | **0.006** | **6.797** | **1.719-26.874** |
| Age of first METH use | 0.152 | 0.304 | 1.165 | 0.871-1.557 |
| Residential district | 0.613 | 0.400 | 1.846 | 0.443-7.694 |
| Alcohol use | 0.164 | 0.723 | 1.179 | 0.474-2.928 |
| Nicotine use | -0.832 | 0.263 | 0.435 | 0.101-1.869 |
| **Heroin use** | **-5.298** | **0.025** | **0.005** | **0.000-0.520** |
| Cannabis use | 0.721 | 0.383 | 2.057 | 0.407-10.404 |
| **Pregabalin use** | **-2.475** | **0.011** | **0.084** | **0.013-0.561** |
| Tramadol use | 0.609 | 0.229 | 1.838 | 0.682-4.952 |
| Corex D use | 1.006 | 0.301 | 2.734 | 0.406-18.386 |
| Thool/Mava use | 0.118 | 0.823 | 1.126 | 0.400-3.165 |
| LSD use | -0.849 | 0.465 | 0.428 | 0.044-4.162 |
| Ecstasy use | 0.034 | 0.984 | 1.035 | 0.035-30.225 |
| Benzos use | 1.714 | 0.088 | 5.549 | 0.774-39.764 |
| Benzhexol use | -0.831 | 0.438 | 0.436 | 0.053-3.558 |

## Discussion

In this study, a reported majority (60%) of methamphetamine users were single males with mean age of 31.30 years. These findings are consistent with similar studies conducted in Sri Lanka, which shows 60.5% being single and 98.5% being males (13). Approximately half of methamphetamine users were reported to have a history of conflict with the law enforcement authorities, which aligns with increased methamphetamine and other substance-related imprisonments noted in recent studies. A total of 13,720 persons were arrested by law enforcement agencies in 2021 for methamphetamine related offences, and its use has risen considerably in the country during the period of 2021 and 2022(7). A family history of substance use was present among 9.8% of users, which is lower compared to similar studies in China, with methamphetamine dependence was at 18.79% and methamphetamine /heroin co-dependence was at 21.05% (14), as well as in Iran, where it was 77.2%(15). The majority of users’ first introducer was a friend, and most had used methamphetamine at a friend’s place or their own home (6). Most of these findings are comparable to those of this study. No participant reported intravenous methamphetamine use, and 8 (3.3%) users indicated they started methamphetamine at night gatherings at funeral houses, which is a specific finding of this study. The rate of methamphetamine dependence among users in a study conducted in Australia(16) was similar to this study, but that study employed the Severity of Dependence Scale in comparison to DSM-5 criteria. The most common comorbid substances were tobacco (33.97%), and alcohol (18.76%), followed by heroin (14.25%) (17). This study identified three common comorbid substances among methamphetamine users: tobacco, heroin, and cannabis, respectively. A significant proportion of participants met the criteria for methamphetamine and heroin co-dependence, although heroin use was not assessed using standard diagnostic criteria for dependency. This may be due to their commonly held belief that methamphetamine act as an opioid substitute to counteract the effects of heroin(18).

Methamphetamine-associated psychosis revealed a unique pattern of psychiatric symptoms in our study. The most common mental illnesses were substance-induced psychosis (10.6%), depression and schizophrenia, and these findings were similar to those of other studies (5). Another study, with using only 19 individuals and DSM-5 diagnostic criteria, revealed schizophreniform disorder, schizoaffective disorder, bipolar affective disorder and unspecified psychotic disorder as the most common psychiatric illnesses among methamphetamine users (19). A Chinese study found that both methamphetamine-dependents and methamphetamine-heroin co-dependent groups had a higher prevalence of current and lifetime risks of developing psychotic disorder (14). In this study, schizophrenia was diagnosed in only 3.7% of consumers. In one study using several scales, including PANSS, the prevalence of methamphetamine-associated psychotic symptoms was 17%, and the top three psychotic symptoms were somatic concerns (36.7%), guilt feeling (93, 36.7%) and excitement (33.9%) (17). In this study, the most common psychiatric symptoms were suspiciousness, hostility, and depressed mood. Another study done in China using several scales, revealed that 57.6% of dependent individuals had any type of psychiatric symptoms including depression, anxiety and psychotic symptoms(20). In the same study, a dose-response relationship was found, and psychiatric symptoms were more common. Psychotic symptoms were associated with methamphetamine dependency and increased duration of use (20).

In this study, 86 participants (36%) had above-average BPRS scores, which were significantly associated with the presence of dependence and methamphetamine use for more than 90 days. The association of psychosis with methamphetamine use within the last 7 days may be due to transient psychotic symptoms in a proportion of methamphetamine users, which may not constitute a psychotic disorder(21). In methamphetamine induced psychotic disorder, symptoms must last beyond the expected effects of intoxication or withdrawal(21). Ideally, methamphetamine users in this study needs to be followed up to determine the course of their illness and for retrospective diagnosis. Logistic regression revealed that heroin use, pregabalin use, and marital status were significant predictors of above-average BPRS scores among methamphetamine users, with odds ratios of 0.005, 0.008, and 0.015, respectively.

This study was conducted at a substance use disorder management clinic in the Western Province of Sri Lanka during the COVID-19 pandemic, when the number of methamphetamine users presenting was lower than expected. As such, the sample may not adequately represent users at the community level. The use of convenience sampling introduces selection bias and limits the generalizability of the findings. Reliance on self-reported information, absence of objective assessments, and the potential influence of other psychoactive substances may have affected the accuracy of psychiatric symptom assessment. In addition, the use of varying street names for substances could have introduced misclassification. Finally, the cross-sectional design restricts causal inference between methamphetamine use and psychosis as well as duration of psychotic symptoms.

Methamphetamine users in Sri Lanka are predominantly young men with high rates of dependence, polysubstance use, and psychiatric morbidity. Dependence was independently associated with higher psychiatric symptom burden. These findings underscore the urgent need for early detection, integrated treatment approaches, and targeted prevention strategies to mitigate the growing public health impact of methamphetamine use in Sri Lanka.

## Data Availability

All data produced in the present study are available upon reasonable request to the authors

## Acknowledgements

We would like to thank the staff of the Substance Use Disorder Clinic of NHSL, Dr. Sumal Nandasena (Managing Director of the Department of Health for Geographic Information System Mapping), and Dr. A. Pubudu De Silva for their statistical support.

## Appendices

## References

1. World Drug Report 2016 [Internet]. [cited 2023 Apr 9]. Available from: https://www.unodc.org/wdr2016/

2. Bramness JG, Rognli EB. Psychosis induced by amphetamines. Current Opinion in Psychiatry. 2016 July 1;29(4):236–41.

3. McKetin R, Lubman DI, Najman JM, Dawe S, Butterworth P, Baker AL. Does methamphetamine use increase violent behaviour? Evidence from a prospective longitudinal study. Addiction. 2014 May;109(5):798–806.

4. Glasner-Edwards S, Mooney LJ, Marinelli-Casey P, Hillhouse M, Ang A, Rawson RA. Psychopathology in methamphetamine-dependent adults 3 years after treatment. Drug Alcohol Rev. 2010 Jan;29(1):12–20.

5. Hides L, Chan G, Dawe S, McKetin R, Kavanagh DJ, Young RM, et al. Direction of the relationship between methamphetamine use and positive psychotic symptoms in regular methamphetamine users: evidence from a prospective cohort study. Br J Psychiatry. 2021 July;219(1):361–7.

6. Research section, National Dangerous Drug Control Board, Sri Lanka. Survey on Methamphetamine(ice) use related prevalences in Sri Lanka 2020. 2020.

7. Reserach division, National dangerous drugs control board, Ministry of public security, Sri Lanka. Handbook of drug abuse information in Sri Lanka 2022.

8. Arunogiri S, Petrie M, Sharkey M, Lubman DI. Key differences in treatment-seeking stimulant users attending a specialised treatment service: a means of early intervention? Australas Psychiatry. 2017 June;25(3):246–9.

9. McKetin R, Lubman DI, Baker AL, Dawe S, Ali RL. Dose-related psychotic symptoms in chronic methamphetamine users: evidence from a prospective longitudinal study. JAMA Psychiatry. 2013 Mar;70(3):319–24.

10. Zanello A, Berthoud L, Ventura J, Merlo MCG. The Brief Psychiatric Rating Scale (version 4.0) factorial structure and its sensitivity in the treatment of outpatients with unipolar depression. Psychiatry Res. 2013 Dec 15;210(2):626–33.

11. Chon Park Y, Kanba S, Chong MY, Tripathi A, Kallivayalil RA, Avasthi A, et al. To use the brief psychiatric rating scale to detect disorganized speech in schizophrenia: Findings from the REAP-AP study. The Kaohsiung Journal of Medical Sciences. 2018 Feb 1;34(2):113–9.

12. Hofmann AB, Schmid HM, Jabat M, Brackmann N, Noboa V, Bobes J, et al. Utility and validity of the Brief Psychiatric Rating Scale (BPRS) as a transdiagnostic scale. Psychiatry Res. 2022 Aug;314:114659.

13. Jayamaha AR, Dharmarathna ND, Herath ND, Ranadeva ND, Fernando MM, Samarasinghe KL, et al. The Pattern of Substance Use and Characteristics of the Individuals Enrolled in Residential Treatment at Selected Rehabilitation Centers in Sri Lanka: A Descriptive Cross-Sectional Study. Subst Abuse. 2022;16:11782218221100823.

14. Dong H, Yang M, Liu L, Zhang C, Liu M, Shen Y, et al. Comparison of demographic characteristics and psychiatric comorbidity among methamphetamine-, heroin- and methamphetamine-heroin codependent males in Hunan, China. BMC Psychiatry. 2017 Dec;17(1):183.

15. Sarani EM, Ahmadi J, Oji B, Mahi-Birjand M, Bagheri N, Bazrafshan A, et al. Investigating the sequential patterns of methamphetamine use initiation in Iran. Substance Abuse Treatment, Prevention, and Policy. 2020 July 29;15(1):52.

16. Quinn B, Ward B, Agius PA, Jenkinson R, Hickman M, Sutton K, et al. A prospective cohort of people who use methamphetamine in Melbourne and non-metropolitan Victoria, Australia: Baseline characteristics and correlates of methamphetamine dependence. Drug Alcohol Rev. 2021 Nov;40(7):1239–48.

17. Su MF, Liu MX, Li JQ, Lappin JM, Li SX, Wu P, et al. Epidemiological Characteristics and Risk Factors of Methamphetamine-Associated Psychotic Symptoms. Front Psychiatry. 2018;9:489.

18. Ellis MS, Kasper ZA, Cicero TJ. Twin epidemics: The surging rise of methamphetamine use in chronic opioid users. Drug Alcohol Depend. 2018 Dec 1;193:14–20.

19. McInnis P, Lee A. Methamphetamine use in an early psychosis service: a crosssectional retrospective cohort study. Australas Psychiatry. 2019 Aug;27(4):383–7.

20. Ma J, Sun XJ, Wang RJ, Wang TY, Su MF, Liu MX, et al. Profile of psychiatric symptoms in methamphetamine users in China: Greater risk of psychiatric symptoms with a longer duration of use. Psychiatry Research. 2018 Apr;262:184–92.

21. Glasner-Edwards S, Mooney LJ. Methamphetamine Psychosis: Epidemiology and Management. CNS Drugs. 2014 Dec;28(12):1115–26.

